# serocalculator, an R package for estimating seroincidence from cross-sectional serological data

**DOI:** 10.1101/2025.06.04.25328941

**Authors:** Kristina W. Lai, Chris Orwa, Jessica C. Seidman, Denise O. Garrett, Samir K. Saha, Dipesh Tamrakar, Farah Naz Qamar, Richelle C. Charles, Jason R. Andrews, Peter Teunis, Kristen Aiemjoy, Douglas Ezra Morrison

## Abstract

**Motivation:** Seroincidence—the rate of new infections in a population—is a key measure for understanding pathogen transmission dynamics and informing public health action, particularly when clinical surveillance is not feasible or reliable for population-level incidence estimation.

**Implementation:** **serocalculator** is an open-source R package that uses a likelihood-based framework incorporating modeled antibody decay, biological variability, and measurement noise to estimate seroincidence rates under Poisson infection processes from cross-sectional serological data.

**General features:** The package supports overall and stratified seroincidence estimation using single or multiple biomarkers. It requires three inputs: (1) a pre-estimated seroresponse model characterizing post-infection antibody waning; (2) noise parameters capturing biological and assay-related variability; and (3) quantitative antibody responses from a cross-sectional serosurvey. It is computationally efficient, well-documented, and includes a point-and-click R Shiny interface. These features promote usability across research and public health.

**Availability:** The package **serocalculator** is freely available on <u>CRAN</u>, with development versions on <u>GitHub</u>.

## Introduction

Serological surveys (serosurveys) can be used to measure population-level antibody responses and characterize infection occurrence (1,2). Following pathogen exposure, the adaptive immune system generates antigen-specific antibodies, which increase rapidly after infection and wane over time (1,3,4). The antibody concentration against a given pathogen reflects the time since last infection, conditional on host, assay, and exposure history (2–4). In cross-sectional serosurveys, a large proportion of individuals with high antibody concentrations suggests high burden of recent or frequent infection, while a smaller proportion indicates lower burden or less recent infection.

Population-level antibody responses are typically characterized using two key epidemiological measures: seroprevalence, the proportion of individuals with antibody levels above a defined threshold; and seroincidence, the rate of new infections in the population (1,2). Here, we define infection as exposure to a pathogen sufficient to induce an immune response, regardless of clinical symptoms. The magnitude and speed of an individual’s immune response to infection is shaped by many factors including infectious dose, age, disease severity, prior exposures, and vaccination history. Seroprevalence-based methods dichotomize quantitative antibody levels into binary outcomes—seropositive or seronegative. While simple and widely used, this approach ignores temporal changes in antibody concentrations and individual-level heterogeneity. Moreover, seroprevalence reflects the cumulative history of prior exposures in a population and is influenced by age distribution, making it an imperfect proxy for recent infection. There is growing interest in methods that leverage quantitative antibody responses and seroincidence estimates to help determine whether a disease has been eliminated (5,6), identify risk factors for infection, and locate areas where rates of new infections are highest (3,7).

New methods estimate seroincidence rates from cross-sectional serosurveys by incorporating models of antibody decay dynamics from patients with confirmed infections (seroresponse parameters) (2,8,9). In brief, this approach uses maximum likelihood estimation (MLE) to determine the most likely seroincidence rate given the observed cross-sectional population data, the known antibody decay trajectories, non-specific binding of the assay probes, and laboratory assay variability (1,2,8–11). This method has previously been used to estimate the seroincidence of enteric fever, a systemic bacterial infection caused by *Salmonella enterica* serovars Typhi and Paratyphi. The study found that while seroincidence estimates were substantially higher than blood-culture-based clinical incidence, they tracked the same rank order of clinical burden across sites (12).

Here, we introduce **serocalculator**, a statistical software package that estimates seroincidence rates from cross-sectional serosurveys by integrating independent models of post-infection antibody kinetics with measurement noise correction (1,8–10). A detailed list of requirements for the **serocalculator** approach are described in Supplement S1. An earlier package, *seroincidence* (13), was developed and maintained from 2015-2018, but lacked methods for incorporating biologic and measurement noise. **serocalculator** supersedes this package by incorporating recent methodological advancements in seroincidence estimation, including biologic and measurement noise and the combination of information across multiple antibody responses and isotypes, while adding functions for simulating cross-sectional data and modern *ggplot2*-based visualizations (14).

The approach requires longitudinal seroresponse data from confirmed infections, assay concordance between the longitudinal and cross-sectional datasets, and quantitative antibody measurements (Supplement S1). The model assumes a constant, homogeneous force of infection (FOI) (Poisson process) with no durable post-infection immunity, generalizability of longitudinal seroresponse parameters to the cross-sectionally surveyed target population, and conditional independence of antibody isotype responses, among other assumptions detailed in Supplement S2 and described previously (1,9,11,15).

### Implementation

**serocalculator** can be downloaded from the <u>Comprehensive R Archive Network (CRAN)</u>. Over 30 exported functions guide users through data import, cleaning, analysis, and visualization. All functions in **serocalculator** version 1.4.1 are described on the <u>package website</u>. The general workflow is: 1) import data, 2) inspect and visualize data, and 3) estimate seroincidence rate. The full process is depicted in Figure 1.

**Figure 1:**
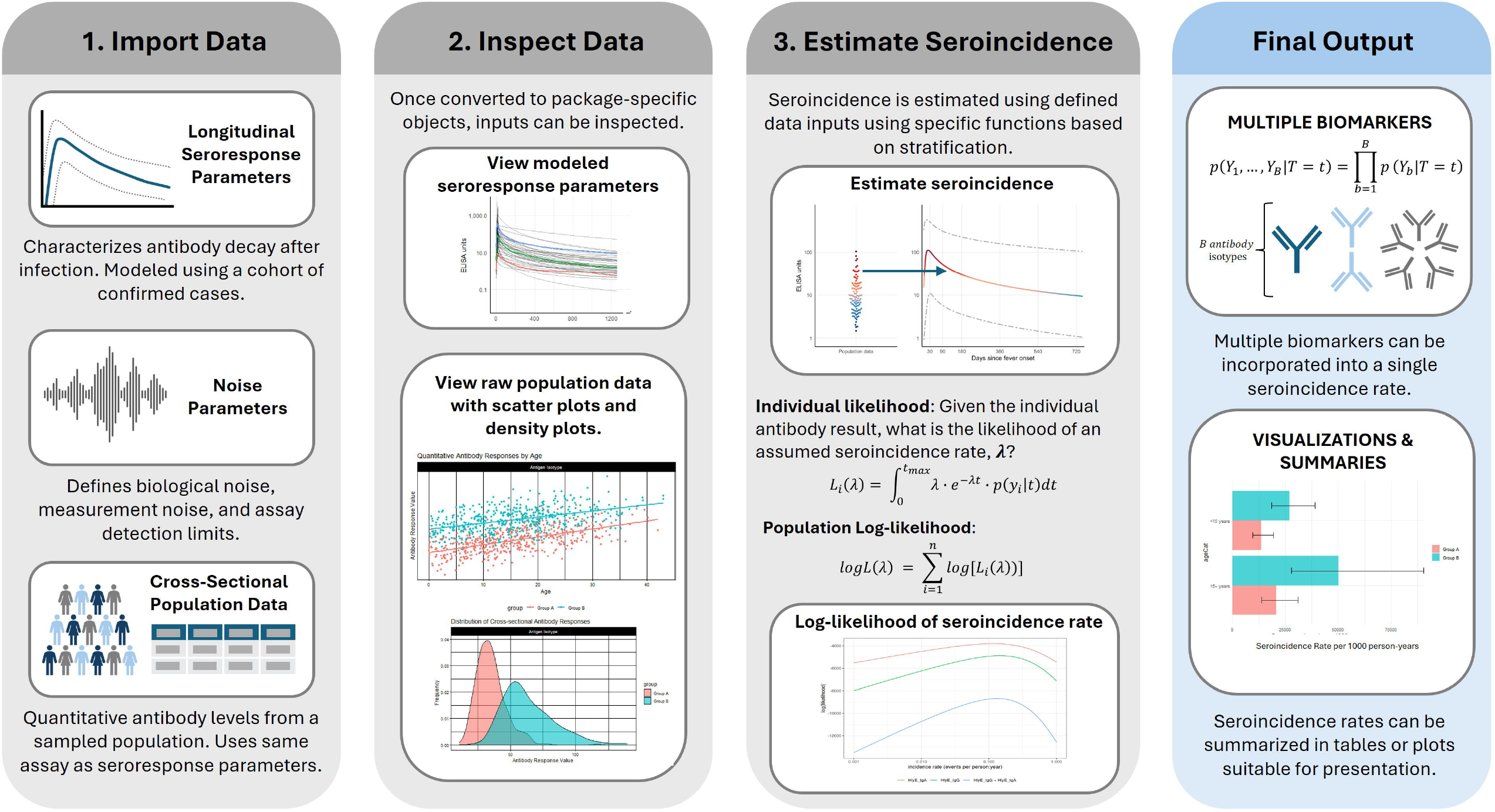
**serocalculator** workflow schematic following 3 steps: 1) import data, 2) inspect data, 3) estimate seroincidence. Final output can be summarized in a table or plot. **Alt Text:** Four panels depicting the general workflow and key equations for **serocalculator**.

### Import Data

Three inputs are required: 1) Previously-estimated seroresponse parameters characterizing longitudinal responses of the surveyed serum antibodies; 2) Noise parameters encompassing biological noise (individual variation in response) and measurement noise (laboratory assay variability); and 3) Quantitative antibody levels from a cross-sectional population-based serosurvey. Users can access small example files within the package, or import larger sample files from the publicly available <u>Serocalculator Data Repository</u> on Open Science Framework. All inputs must use the same assay and calibration to be comparable. Further requirements, formatting guidance, and example input datasets are provided in Supplements S1 and S3.

#### Longitudinal Seroresponse Parameters

Longitudinal seroresponse parameters describe how antibodies rise and fall after infection. Before using **serocalculator** to estimate seroincidence, these parameters must be estimated by fitting two-phase, hierarchical within-host models to quantitative antibody responses measured longitudinally in a cohort of confirmed cases, using Bayesian inference with Markov chain Monte

Carlo (MCMC) to sample from the joint posterior distribution of the model parameters for each antigen-isotype (1,8–10). A companion package, *serodynamics*, expands **serocalculator**’s applicability across diverse research settings by enabling users to model custom seroresponse parameters for additional pathogens and quantitative assays from their own longitudinal data (16). Further details can be found in Supplement S4.

Users can import seroresponse parameters in CSV or RDS format, then apply `as_sr_params()` to assign the imported dataset as an `sr_params` object (a subclass of `tibble::tbl_df` with added metadata attributes) and ensure required variables are present and in the correct format. If using files from a URL, users can apply `load_sr_params()` to properly format data. The required seroresponse parameter variables are: one or more disease-specific antigen and antibody isotype pairs matching those in the cross-sectional population data (`antigen_iso`), baseline antibody concentration (`y0`), peak antibody concentration (`y1`), time to peak antibody concentration (`t1`), antibody decay rate (`alpha`), and antibody decay shape (`r`).

#### Noise Parameters

The noise parameters object defines assumptions about the measurement process (1). Biological noise (*v*) reflects potential non-specific antibody binding from other exposures, such as cross-reactivity with a related pathogen. It is estimated by the 95th percentile of a distribution of antibody responses to the antigen-isotype in a population with low or no prior exposure to the pathogen. Measurement noise (ϵ) represents laboratory assay variability from the laboratory testing process. It is defined by a coefficient of variation (CV), or the ratio of the standard deviation to the mean for replicates across plates. **serocalculator** can also accommodate censored data: lower and upper limits of reliable quantification may be specified as y_low_ and y_high_, respectively, and observations outside of those limits will be censored. Further details on noise parameters are in Supplement S5.

As above, users can import noise parameters (CSV or RDS) as a `data.frame,` then apply `as_noise_params()`, to get a correctly formatted `noise_params` object. If using files from a URL, users can apply `load_noise_params()` and follow the process described above for the seroresponse parameters. The required noise parameter variables are: one or more disease-specific antigen and antibody isotype matching those in the seroresponse parameters (`antigen_iso)`, biological noise (`nu`), measurement noise (`eps`), and lower and upper limits of detection of the assay (`y.low` and `y.high,` respectively).

#### Cross-sectional population data

Once again, users can import cross-sectional population data in either CSV or RDS format and apply `as_pop_data()`, which assigns the imported dataset as a `pop_data` object, ensuring correct formatting. The required variables for this dataset are: age in years (`age`), one or more specified antigen isotype pairs (`antigen_iso)`, quantitative antibody response (`value`), and unique participant id (`id`). Additional variables can be included for stratification. The `as_pop_data()` function detects the required variables or allows users to specify them. If using data from a URL, users can apply `load_pop_data()` and then once again follow the process described above.

### Estimating seroincidence

Once inputs are imported and formatted, users can estimate overall and stratified seroincidence rates using `est_seroincidence()` and `est_seroincidence_by()`, respectively. Users specify the three required data inputs and select one or more antigen isotype pairs for estimation.

Optional parameters allow customization of visualizations, warning messages, parallel computing, and the estimation process. Users can generate stratified seroincidence estimates by specifying strata variables in the cross-sectional dataset.

### Use

We will demonstrate the use of **serocalculator** with a reproducible example of enteric fever seroincidence using a subset of data from serosurveys conducted in Bangladesh, Nepal and Pakistan in 2016-2021 (12,17). Additional details are available in Supplement S6 and data are publicly available in the <u>Serocalculator Data Repository</u> on Open Science Framework (18). This example is also presented as an <u>article</u> on the **serocalculator** <u>website</u>. Code and output for this example are provided in Figure 2.

**Figure 2:**
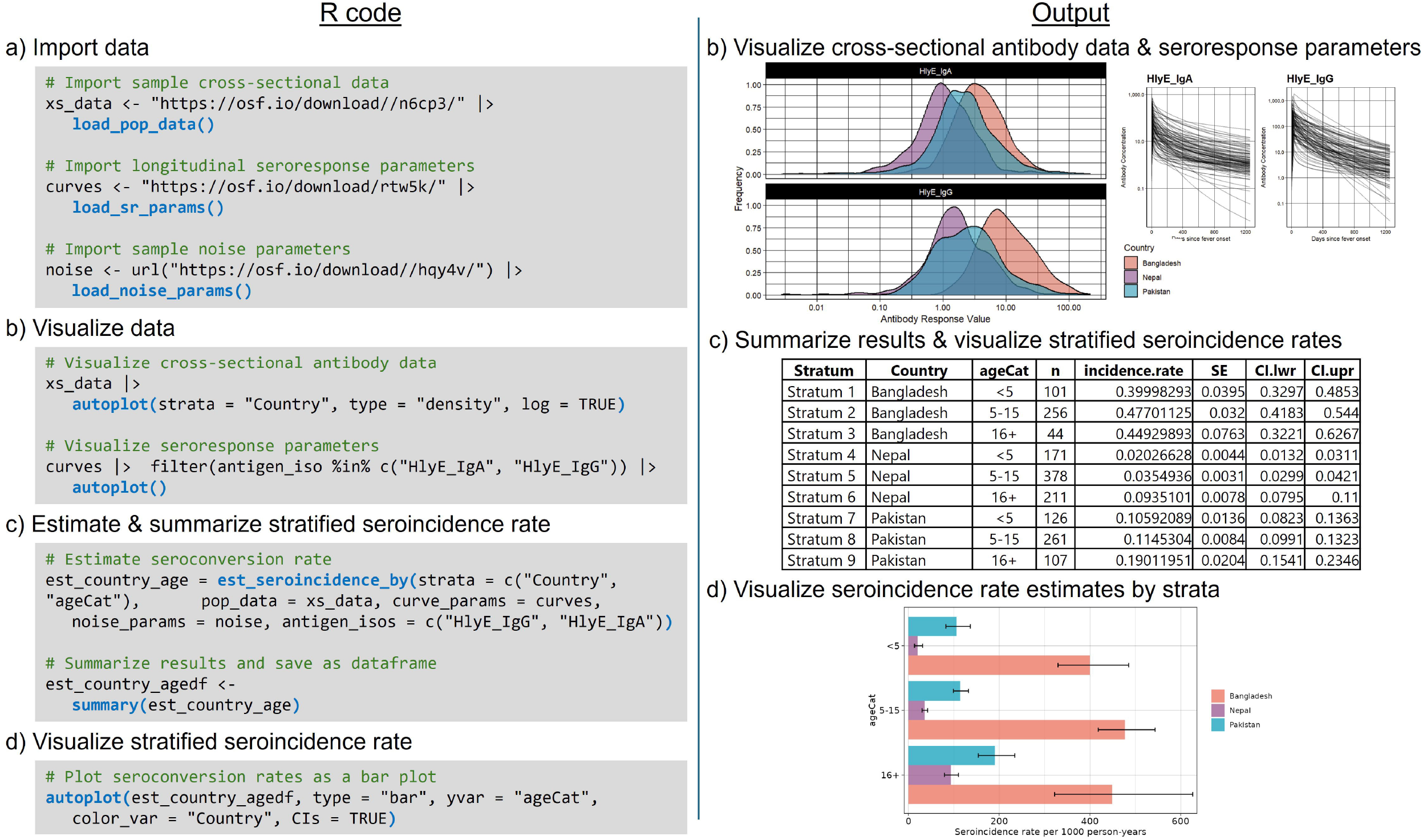
Estimation of seroincidence rates stratified by country and age category. a) Code for importing the three required data inputs. b) Code and output for visualizing cross-sectional antibody data and longitudinal seroresponse. c) Code and output for executing and summarizing results of the `est_seroincidence_by()` function. d) Code and output for visualizing stratified seroincidence results as a bar plot. **Alt Text:** R code and output showing steps of data import, seroincidence estimation, results summary, and visualization in subfigures labeled from a to d.

First, we load each of our three inputs (seroresponse parameters, noise parameters, and cross-sectional population data) and then use built-in `autoplot()` visualization methods to create descriptive plots to inspect cross-sectional data and modeled seroresponse parameters. In this example, we import noise parameters calculated at each study site laboratory from analysis using enzyme-linked immunosorbent assays (ELISA).

We estimate overall and stratified seroincidence rates using the `est_seroincidence()` and `est_seroincidence_by()` functions, respectively. In both cases, we combine IgA and IgG seroresponses to the hemolysin E (HlyE) antigen. For the overall seroincidence rate, we filter to only Pakistan and find a seroincidence rate of 128 new infections per 1000 person-years (95% CI: 115-142). To compare across countries and ages, we specify the strata variables and find that 5-to-15-year-olds in Bangladesh experience the highest seroincidence of enteric fever, with a rate of 477 new infections per 1000 person-years (95% CI: 418-544). Model outputs from `est_seroincidence()` and `est_seroincidence_by()` can be summarized and visualized using the corresponding `summary()` and `autoplot()` methods (Figure 2).

## Discussion

**serocalculator** provides a computationally efficient method for calculating population-level seroincidence rates from multiple biomarkers while accounting for antibody decay dynamics, biologic variability, and measurement noise. By updating analytic methods from its predecessor

(13) and integrating new tutorials, simulations, and visualizations, the package significantly expands the utility and accessibility of quantitative seroepidemiology. Thus far, **serocalculator** has been applied to enteric fever and scrub typhus studies (12,19–21), and its methods are increasingly adopted by research teams and public health institutions to inform typhoid vaccine introduction and surveillance in endemic regions.

Several groups have developed R packages to support seroepidemiologic analyses. For cross-sectional data, *serosv* estimates seroprevalence and force of infection (FOI) using a wide array of parametric and non-parametric models applied to cross-sectional serosurvey data (22). Similarly, *serofoi* employs serocatalytic models to estimate FOI using constant, age- and time-varying models (23). Extending these cross-sectional approaches, *Rsero* provides Bayesian tools for comparing serocatalytic models, assessing FOI under various assumptions (e.g., transmission mode, seroreversion), and evaluating sampling strategies, though it relies on binary serostatus (24). *serosim* simulates serological data to support study design and evaluate within-host dynamics and new analytic approaches (25). Like *serosim* and **serocalculator**, *serojump* and *serosolver* leverage quantitative antibody kinetics rather than binary classification. *serojump* uses individual-level cohort data to focus on individual infection timing and antibody kinetics using Bayesian reversible jump MCMC (26), while *serosolver* utilizes a hierarchical Bayesian framework to reconstruct infection histories for complex, cross-reactive multi-strain pathogens (27).

In contrast, **serocalculator** is explicitly designed to estimate population-level seroincidence rates by pairing cross-sectional quantitative antibody responses with modeled antibody decay curves, accounting for individual-level heterogeneity, multiple antigen isotypes, and both within-individual measurement error and between-individual biologic variability. Collectively, these packages expand access to seroepidemiologic tools, with **serocalculator** filling a distinct methodological niche for cross-sectional serosurveillance. Future work will assess the impact of simplifying computational assumptions and explore structural modifications to relax them.

Current limitations present opportunities for methodological improvement. First, ignoring frequent repeated exposures may bias seroincidence estimates in high-burden settings by altering baseline longitudinal seroresponse distributions (11). Because the current MLE approach does not address this, seroincidence estimates should be interpreted with caution in high-burden settings where repeated exposure is likely, as prior immunity and boosting may skew results depending on the degree of immune memory, exposure interval, and antigen isotype. We are actively working to incorporate simulation-based inference to improve estimation accuracy under high FOI (11). Second, while vaccine and surveillance antigens differed in our enteric fever use cases, future work should integrate vaccination status for pathogens where natural and vaccine-induced responses overlap. Finally, relaxing the assumption of constant infection pressure to account for seasonality or other temporal variation remains an area for future research.

Additional documentation and tutorials for **serocalculator** are available at https://ucd-serg.github.io/serocalculator/.

## Data Availability

All data included and produced are available online at https://osf.io/ne8pc/files/osfstorage and https://github.com/UCD-SERG/serocalculator

https://osf.io/ne8pc/files/osfstorage

## Declarations

### Use of Artificial Intelligence (AI)

The authors utilized Gemini 3.5 Flash for text editing (i.e. to improve readability, clarify grammar, and reduce wordcount) during the manuscript preparation and take full responsibility for the accuracy and integrity of the published content.

### Supplementary data

Supplementary material are available online.

### Data availability

Source code for the **serocalculator** package is available on GitHub: https://github.com/UCD-SERG/serocalculator. Deidentified data from the sample analysis is hosted on Open Science Framework: https://osf.io/ne8pc/files/osfstorage.

