## Supplement for "serocalculator, an R package for estimating seroincidence from cross-sectional serological data"

### S1. Methodological Requirements

These are prerequisites that must be satisfied before the serocalculator framework can be validly applied. They are largely within the investigator's control through study design and data preparation.

| Requirement | Description | Consequence if Not Met |
|---|---|---|
| **1. Availability of Longitudinal Seroresponse Data** | The *serocalculator* package requires longitudinal antibody measurements from a cohort of individuals with confirmed infections to estimate seroresponse parameters ($y_0$, $y_1$, $t_1$, $\alpha$, $r$).[1,2] These parameters define the expected antibody trajectory following infection and serve as fixed inputs to the cross-sectional seroincidence model. | Without longitudinal seroresponse data, the model has no basis for interpreting antibody levels in the cross-sectional population. The framework cannot be applied. |
| **2. Assay Concordance Between Longitudinal and Cross-Sectional Data** | The longitudinal cohort and the cross-sectional survey must use the same serological assay, antigen targets, measurement units, and normalization procedures. The seroresponse parameters are calibrated to a specific assay's dynamic range and scale.[3] | If assays differ (e.g., ELISA in EU/mL vs. Luminex in MFI), the seroresponse model cannot accurately map observed antibody levels to time since infection. Estimates would be uninterpretable. |
| **3. Quantitative Antibody Measurements** | The method requires quantitative (continuous) antibody concentrations rather than binary (positive/negative) or qualitative results. The full distribution of antibody levels in the cross-sectional sample is used to construct the likelihood.[1,4] | Binary seroclassification discards the quantitative information essential for estimating time since infection.[2] Dichotomized data would preclude use of the serocalculator framework entirely. Assays must provide reliable quantitative output across the relevant dynamic range of antibody concentrations. |
| **4. Age Data Availability** | The model requires participant age to correct for the age-dependent probability of never having been infected. This correction was introduced by Teunis & van Eijkeren (2020).[2] | The present implementation of serocalculator cannot compute seroincidence rates if age data is not provided. |
| **5. Selection of Infection-Specific Antigen Targets** | Antigen targets used in the serological assay should be specific to the pathogen of interest and should not react with closely related pathogens likely to elicit cross-reactive responses. | If the target antigen is shared with a related pathogen, the model cannot distinguish infection from the pathogen of interest with that of a cross-reactive pathogen. This can be mitigated by selecting pathogen-specific antigen targets (e.g., hemolysin E for typhoid).[5] |

| Requirement | Description | Consequence if Not Met |
|---|---|---|
| **6. Exclusion of Vaccine-Related Antigen Targets** | Antigen targets used in the serological assay should not be shared with vaccines in use in the target population and should not cross-react with vaccine-induced responses. | If the target antigen is shared with a vaccine that has been used in the population (e.g., Vi polysaccharide for typhoid) the model cannot distinguish infection-induced from vaccine-induced antibody responses. This can be mitigated by selecting infection-specific antigen targets (e.g., hemolysin E for typhoid).[5] |

### S2. Modeling Assumptions

These are simplifying assumptions embedded in the statistical and biological model. They are not fully within the investigator's control and may or may not hold in practice.

| Assumption | Description | Implications of Violation |
|---|---|---|
| **1. Generalizability of Seroresponse Parameters** | The seroresponse parameters modeled from a longitudinal cohort of confirmed infections are assumed to be representative of the antibody kinetics in the broader population and therefore generalizable to the target cross-sectional population.[1,2] The distributions of peak antibody levels and decay rates estimated from this cohort are treated as generalizable to the broader population of infections, including subclinical and mild cases. | If the longitudinal cohort is not representative — for example, due to spectrum bias (symptomatic/culture-confirmed cases mounting stronger immune responses than subclinical infections), mismatched age groups, differing levels of immunosuppression, or prior exposure history — seroincidence estimates may be biased. Specifically, if the cohort overrepresents severe cases with higher peak antibody levels, the model may overestimate time since infection and thus underestimate seroincidence.[3,4] |
| **2. Constant Force of Infection (Poisson Process)** | Infections are modeled as a homogeneous Poisson process, meaning all individuals in the population are subject to a constant and time-invariant seroconversion rate ($\lambda$).[1,5] The time between successive infections is exponentially distributed. Infection risk does not vary across individuals or subgroups unless rates are explicitly stratified by those factors. | If the true force of infection varies over time (e.g., due to seasonality, epidemic waves, or secular trends), the seroincidence estimate reflects a time-averaged rate that may not represent current transmission intensity.[5] Estimates may under- or overestimate rates depending on whether samples are collected during low or high transmission periods. This limitation can be partially mitigated by sampling consistently over time or stratifying estimates by time period. |
| **3. No Durable Post-Infection Immunity** | Individuals are assumed to return to a fully susceptible state immediately following infection, such that prior infection does not reduce the hazard of reinfection. Each individual faces the same constant force of | If previously infected (or vaccinated) individuals acquire durable protective immunity that reduces susceptibility to reinfection, they would no longer be part of the at-risk population. The model would apply the population-level force of infection |

| Assumption | Description | Implications of Violation |
|---|---|---|
| | infection regardless of infection history.[1,5] | to protected individuals, leading to overestimation of seroincidence. This assumption is most defensible for pathogens where natural immunity is incomplete or short-lived (e.g., many enteric pathogens) but may be substantially violated for pathogens conferring sterilizing immunity.[5] |
| **4. Conditional Independence of Antibody Isotypes/Antigens** | Antibody responses from each isotype (e.g., IgG, IgM, IgA) and antigen are assumed to be conditionally independent given the time since infection (*T*).[1,2] Conditional on a shared infection time, the response to one isotype does not inform or depend on the response to another, because they reflect distinct immunological processes with different kinetic profiles. This allows the joint likelihood to be computed as the product of isotype-specific likelihoods, increasing precision. | If antibody isotype responses are correlated beyond what is explained by shared infection timing (e.g., through shared immunological regulation), treating them as independent overestimates the precision of the seroincidence estimate (i.e., standard errors will be artificially narrow) because each isotype is counted as contributing fully independent information. The effect on the point estimate of λ is uncertain and depends on the unknown correlation structure; bias may or may not result. |
| **5. Parametric Antibody Decay Model** | The post-infection antibody trajectory is described by a specific parametric model: a rapid rise to peak followed by monotonic decay. The original 2012 formulation used exponential decay;[1] the 2016 update extended this to power-function decay,[6] interpretable as a mixture of exponential decay curves reflecting within-host heterogeneity in antibody production. The 2020 update incorporated these nonexponential kinetics into the seroincidence estimation framework.[5] Inter-individual | If the true antibody kinetics do not conform to the assumed parametric form, estimates of time since infection — and consequently seroincidence — may be biased. Misspecification of the decay model can lead to systematic bias, particularly at longer times since infection.[6,7] |

| Assumption | Description | Implications of Violation |
|---|---|---|
| | heterogeneity in these parameters is modeled as random effects, typically on the log scale. | |
| **6. Most Recent Infection Dominates the Antibody Signal** | In the standard likelihood-based implementation, the observed antibody level is assumed to be determined solely by the time since the most recent infection. Prior infection history is ignored; antibodies from earlier infections are assumed to have decayed to negligible levels.[1,2] | In high-transmission settings where repeated infections are common, antibody levels may reflect cumulative boosting from multiple infections rather than a single event. The standard likelihood approach has been shown to underestimate the force of infection under these conditions.[8] An approximate Bayesian computation (ABC) extension was developed to account for repeated infections using Kolmogorov deviance as a goodness-of-fit measure, but this is not yet implemented in the present serocalculator package.[8] |
| **7. Representative Cross-Sectional Sampling** | The cross-sectional serum samples should constitute a representative (ideally random or probability-based) sample of the target population. Each individual contributes a single time-point antibody measurement.[1] | Non-representative sampling (e.g., blood donors, pregnant women, residual clinical sera) may bias population-level seroincidence estimates in either direction. |
| **8. Accurate Noise Specification** | The model incorporates noise parameters to account for biological variability (e.g., baseline antibody levels from cross-reactive antigens or nonspecific immune activation) and assay-related measurement error. These noise parameters must be accurately specified.[5] Users are not able to control biologic noise, but they are able to control and directly estimate measurement | If noise is unaccounted for or misspecified, considerable bias can be introduced, especially at low seroincidence rates (<0.1/yr) and in young age groups.[5] Underestimating noise may inflate seroincidence estimates by attributing background antibody levels to recent infection; overestimating noise may attenuate the signal from true infections. The ABC extension by Teunis et al. (2023) facilitates joint estimation of noise parameters alongside seroconversion rates, but this is |

| Assumption | Description | Implications of Violation |
|---|---|---|
| | error during laboratory testing. | not yet implemented in the present serocalculator package.[8] |
| **9. No Differential Mortality** | The model implicitly assumes that infection does not substantially alter survival — there is no differential mortality between infected and uninfected individuals that would deplete the cross-sectional sample of previously infected persons. | For pathogens that cause substantial mortality, the cross-sectional population would be enriched for survivors and uninfected individuals, potentially leading to underestimation of seroincidence because individuals who mounted strong immune responses but subsequently died would be absent from the sample. |
| **10. No Residual Cross-Reactivity** | After appropriate antigen selection (see Requirements, Supplement S1), the model assumes that measured antibody responses are specific to the pathogen of interest and are not meaningfully confounded by residual cross-reactive responses from antigenically related pathogens.[4] | If residual cross-reactivity inflates observed antibody levels despite appropriate antigen selection, the model may attribute these elevated levels to recent infection, leading to overestimation of seroincidence. The magnitude of this bias depends on the degree of antigenic similarity between the target pathogen and co-circulating related organisms. |

**References and Further Reading**

### S3. Dataset Formatting Requirements

The table shells below are examples of formatted input datasets for each of the three required inputs for **serocalculator**. Any of the inputs may include variables to stratify by, but only strata in the cross-sectional population data will produce stratified seroincidence estimates. Noise parameters and longitudinal seroresponse parameters may be used by strata or as overall values applied equally to each cross-sectional strata.

**Table S3.1:** Longitudinal seroresponse parameters

| antigen_iso | y0 | y1 | t1 | alpha | r | strata1 | iter* |
|---|---|---|---|---|---|---|---|
| A | | | | | | X | 1 |
| A | | | | | | Y | 2 |
| B | | | | | | X | 1 |
| B | | | | | | Y | 2 |

- **antigen_iso** = Antigen and antibody isotype pair(s) measured
- **y0** = Baseline antibody concentration (U/mL)
- **y1** = Peak antibody concentration (U/mL)
- **t1** = Time to peak antibody concentration (days)
- **alpha** = Antibody decay rate (1/days)
- **r** = Antibody decay shape
- **strata variables** (optional) = any demographic features or covariate relevant to your analysis (must match or be a subset of the stratifying variables included in the cross-sectional data)
- **iter** *(from seroresponse model)** = MCMC iteration. Rows correspond to posterior draws from the Bayesian seroresponse model. Not specified by the user (see note)

** Note: The longitudinal seroresponse parameter table will typically contain many rows per antigen-isotype and stratum combination corresponding to posterior draws from the Bayesian seroresponse model. Seroincidence functions can be run with a single iteration, but estimates will be less stable and will not reflect uncertainty in the modeled seroresponse parameters.*

**Table S3.2:** Noise parameters

| antigen_iso | y.low | y.high | eps | nu | strata1 |
|---|---|---|---|---|---|
| A | | | | | X |
| A | | | | | Y |
| B | | | | | X |
| B | | | | | Y |

- **antigen_iso** = Antigen and antibody isotype pair(s)

- **y.low** = Lower limit of detection of the assay used to produce the cross-sectional survey data set (a)
- **y.high** = Upper limit of detection of the assay
- **eps** = Measurement noise of the assay; CV%, range 0-1 (See Supplement S6)
- **nu** = Biologic noise of the assay; 95% of the distribution among those never exposed (See Supplement S6)
- **strata variables** (optional) = any demographic features or covariates relevant to your analysis (must match or be a subset of the stratifying variables included in the cross-sectional data)

**Table S3.3** Cross-sectional population data

| id | age | antigen_iso | value | strata1 |
|---|---|---|---|---|
| 1001 | | A | | X |
| 1001 | | B | | X |
| 1002 | | A | | X |
| 1002 | | B | | X |
| 1003 | | A | | Y |
| 1003 | | B | | Y |

- **id** = Unique identifier for each individual
- **age** = Numeric age in years (can include decimals)
- **antigen_iso** = Antigen and antibody isotype pair(s)
- **value** = Quantitative antibody response value
- **strata variables** (optional) = Any demographic features or covariate relevant to your analysis (must match those in other inputs to be used in analysis)

**Sample Size**

We do not currently recommend a specific sample size in the cross-sectional population data because the required sample depends on the research question and heterogeneity in post-infection seroresponse parameters. Users should use particular caution when producing stratified estimates, as small numbers (e.g. n<20) may produce unstable results. At this time, we recommend that users conduct simulation exercises to determine the power that they already have in an existing cross-sectional study dataset with given seroresponse parameters, or the size needed to detect a desired effect size. Users can use the \`sim_pop_data()\` function in *serocalculator* for simulation-based sample size estimation. For further details, see the Simulation Studies article on our website.

### S4. Mathematical Theory

This supplement describes the mathematical framework used in **serocalculator**. This approach estimates seroincidence rates ($\lambda$) from cross-sectional serosurveys by integrating independent models of post-infection antibody kinetics with measurement noise correction to obtain a marginalized likelihood for $\lambda$. Further details on the seroresponse parameters and noise parameters are provided in Supplements S5 and S6, respectively.

#### S4.1 Latent Infection Time

The incidence rate ($\lambda$) is assumed to be constant over the age range and calendar period represented in the cross-sectional serosurvey. [1,2] For a participant of age $a$, the time since their most recent infection ($T$) is a latent, unobserved variable.

Under the assumption of constant and homogeneous incidence, the latent time since most recent infection ($T$) follows an exponential distribution truncated by age ($a$) at the time of observation.

$$p(T = t \mid A = a) = \underbrace{\mathbb{1}_{\{t \in [0,a]\}}\, \lambda e^{-\lambda t}}_{\text{Ever Infected}} + \underbrace{\mathbb{1}_{\{t = \text{NA}\}}\, e^{-\lambda a}}_{\text{Never Infected}}$$

- **Ever Infected**: The first term represents the probability density of a participant being last infected $t$ days ago. It is the product of the probability of being infected at time $t$ (the incidence rate $\lambda$) and the probability of not being infected again between time $t$ and the survey date ($e^{-\lambda t}$).
- **Never Infected:** The second term, $e^{-\lambda a}$, represents the discrete probability that a participant has escaped infection for their entire life up to their current age. In this state, $T$ is undefined (NA).

Explicitly incorporating the "never infected" probability is essential for removing age-dependent bias. Without this term, the model would incorrectly attribute the low antibody titers common in unexposed children to the long-term decay of a past infection, leading to a significant underestimation of the incidence rate. [3]

#### S4.2 The Three-Part Censored Likelihood Partition

To account for the physical constraints of laboratory assays, the likelihood for an observed titer $y$ at age $a$ is defined by a three-part partition based on lower $(y_L)$ and upper $(y_U)$ detection limits:

$$\ell(y|a; \dots) = \begin{cases} R(y_L|a)\ if\ y \leq y_L \\ \rho(y|a)\ if\ y_L < y < y_U \\ 1 - R(y_U|a)\ if\ y \geq y_U \end{cases}$$

This piecewise approach ensures that participants with results outside the quantifiable range are appropriately weighted. Participants below the detection limit—who are often indistinguishable from the background signal—contribute to the likelihood via the cumulative probability $R(y_L|a)$, preventing the bias that would occur if these values were discarded or assigned arbitrary exact values. [3]

#### S4.3 Seroresponse Kinetics and Kinetic Heterogeneity

Linking the observed titer $(y)$ back to the infection time $(t)$ requires a model for the antibody response after infection, $p(Y = y|T = t)$. As detailed in Supplement S1, the response follows a two-phase model. The active infection phase rises exponentially from a baseline $y_0$ to a peak level $y_1$ at time $t_1$. After the pathogen is cleared at time $t_1$, the antibody concentration enters a decay phase governed by a power-law model. The antibody concentration $y(t)$ during the decay phase is defined as:

This decay is governed by the decay rate $(\alpha)$ and the decay shape parameter $(r)$. When $r > 1$, the model accounts for a rapid initial decline followed by a prolonged period of slower decay, effectively representing immunological memory. When $r = 1$, the model reduces to standard exponential decay. [4,5]

The full seroresponse is summarized by the five-parameter kinetic set $\theta = (y_0, y_1, t_1, \alpha, r)$. To account for between-person heterogeneity, the model averages the likelihood over a Monte Carlo sample of $M$ posterior draws $(\theta^{(k)})$ derived from longitudinal confirmed-case data. This marginalization ensures that the final incidence estimate propagates both individual heterogeneity and parameter uncertainty. [2,3,5]

$$p(Y = y|T = t)\ \approx \frac{1}{M}\sum_{k=1}^{M} p(Y = y|T = t, \theta^{(k)})$$

#### S4.4 Full-Sample Marginal Likelihood

Using the Law of Total Probability, the likelihood of an individual's observed titer is obtained by integrating over all possible infection times ($t$) and the discrete case of no prior infection: [2]

$$p(Y = y) = \int_0^a p\,(Y = y|T = t)p_\lambda(T = t)dt$$

The full-sample likelihood for a survey of $n$ individuals is the product of these individual probabilities:

$$\mathcal{L}(\lambda) = \prod_{i=1}^{n} p\,(Y = y_i) \;\rightarrow \log L(\lambda) \;=\; \sum_{i=1}^{n} \log[p(Y = y_i)]$$

The natural log of this likelihood is numerically maximized to find the maximum likelihood estimate (MLE) for $\lambda$.

#### S4.5 Multiple Biomarkers and Conditional Independence

When a survey measures multiple biomarkers (e.g., antibody isotypes IgG and IgA), they are assumed conditionally independent given the shared time since infection ($T$). For $B$ biomarkers, the joint likelihood factors into a product of per-biomarker terms:

$$p(Y_1, \dots, Y_B|T = t) = \prod_{b=1}^{B} p\,(Y_b|T = t)$$

This simplification allows the participant-level log-likelihoods to be summed. Each biomarker retains its own kinetic and noise parameters but shares the same latent $T$ and the incidence rate $\lambda$ being estimated.

#### S4.6 Numerical Optimization and Standard Errors

Because the log-likelihood involves a sum of logs of integrals, it has no closed-form derivative and must be solved by numerical optimization, using a Newton-type algorithm (*stats::nlm()* in the current implementation). Standard errors are estimated from the Hessian matrix (the curvature of the log-likelihood at its maximum), where greater curvature corresponds to a sharper peak and a smaller standard error.

#### Notation

- $\lambda$: Population incidence rate (per person-time).
- $T$: Latent time since last infection ($NA$ if never infected).
- $a$: Age at sampling.
- $y$: Observed antibody titer.
- $R(y|a)$: Cumulative distribution function of the titer (probability at or below y).
- $\rho(y|a)$: Probability density of the titer.
- $y_L, y_U$: Lower and upper assay detection limits.
- B: Number of biomarkers.
- $\theta$: Kinetic parameter set $(y_0, y_1, t_1, \alpha, r)$.
- $M$: Number of Monte Carlo draws for marginalization.

### S5. Further Details on Longitudinal Seroresponse Parameters

The longitudinal seroresponse parameters in Table S1.1 are components of the within-host seroresponse model proposed in de Graaf *et al.* 2014 and extended in Teunis *et al*. 2016 and Diekmann *et al.* 2018.[1-3] The model consists of two phases: an active-infection/antibody growth phase and a post-infection/antibody decay phase. Beginning at time $t_0$, or the initial time of infection, there is an initial exponential growth phase (Figure S5.1a) until the peak antibody response ($y_1$ at time $t_1$). Note that this brief growth phase is presumed to be minimal and is ignored in the final seroincidence model. After $t_1$, we model the antibody decay phase (Figure S5.1b) using a power function, with the parameters listed below:

| | | | |
|---|---|---|---|
| Growth phase: | $y(t) = y_0 e^{\mu t},$ | $[0 \leq t < t_1]$ | Figure S5.1a |
| Decay phase: | $y(t) = y_1\left(1 + (r-1) y_1^{r-1} \alpha (t - t_1)\right)^{-\frac{1}{r-1}},$ | $[t_1 \leq t < \infty]$ | Figure S5.1b |

Parameters:
- $y_0$ = Baseline antibody concentration (U/mL); i.e., $y_0 = y(0)$
- $y_1$ = Peak antibody concentration (U/mL); i.e., $y_1 = y(t_1)$
- $\mu$ = Antibody growth rate
- $t_1$ = Time to peak antibody concentration (days)
- $\alpha$ = Antibody decay rate (1/days)
- $r$ = Antibody decay shape

This model is fit in a Bayesian framework using 'JAGS' and the posterior distributions ($\boldsymbol{y_1}, \boldsymbol{r}, \boldsymbol{\alpha}$) are then used in **serocalculator**. A companion R package, *serodynamics*, is available on CRAN to aid users in modeling post-infection seroresponse parameters from longitudinal cohorts for confirmed cases. These seroresponse models can be stratified by age, country, and/or other relevant demographic characteristics if seroresponse dynamics vary among the corresponding subpopulations.

**Figure S5.1a-b:** Grey dots and lines depict observed longitudinal antibody responses measured from confirmed cases, each dot is a measured value and the lines connect measurements in an individual. The multi-colored line is the median modeled antibody response, red indicates higher quantitative antibody responses and blue indicates lower responses. The blue dashed line indicates time to peak antibody response (y1).

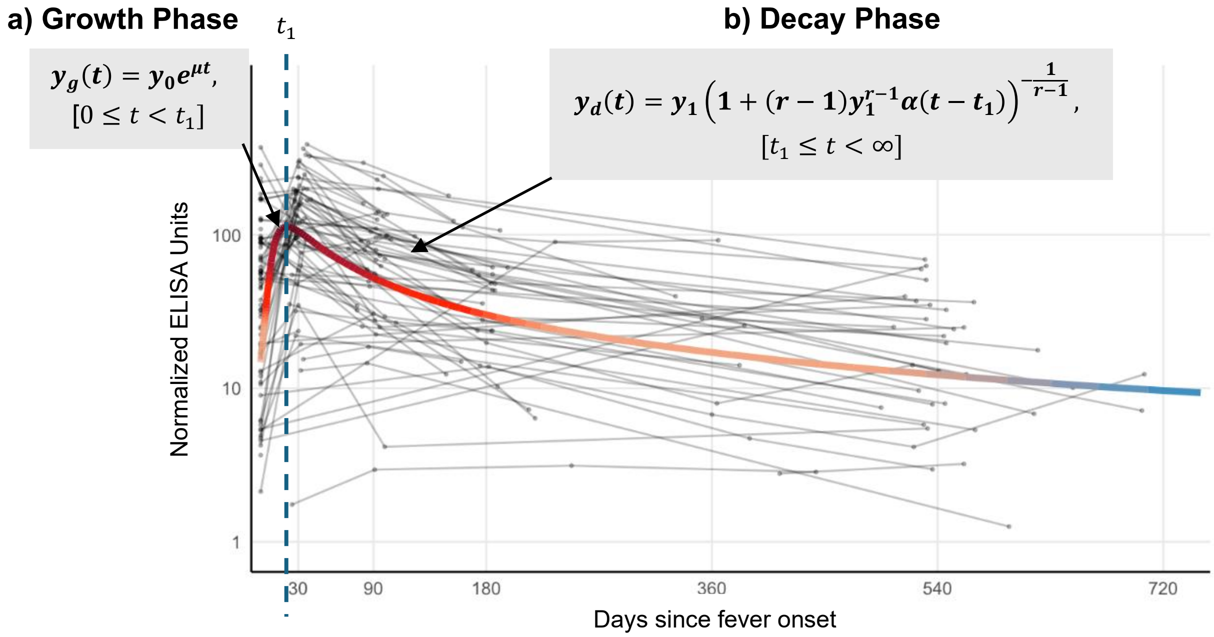

**References and Further Reading:**

### S6. Further Details on Noise Parameters

**serocalculator** is designed to incorporate two sources of noise: biological noise from cross-reactivity or non-specific binding, and measurement noise during laboratory analysis. In the package, biologic noise is denoted by $\nu$ ("nu") and measurement noise is denoted by $\varepsilon$ ("epsilon").[1]

The biological noise, $\nu$, represents error from cross-reactivity to other antibodies. This parameter is estimated by the 95th percentile of a distribution of antibody responses to the antigen-isotype in a reference population with low or no recent exposure to the pathogen of interest. This specific percentile is used because incidence estimates are highly sensitive to the scale of noise but robust to its distributional shape. Simulations have shown that a simple uniform distribution spanning this 95% range accurately approximates more complex noise distributions (e.g., lognormal), ensuring that background variation is correctly attributed to noise rather than interpreted as evidence of infection.[1] Biological noise is assumed to be random and is always positive.

The reference population used to define $\nu$ should represent a population in which recent infection with the pathogen of interest is unlikely, such as data from geographically distinct low-transmission populations or archived pre-epidemic samples. If $\nu$ is underspecified, the model may incorrectly attribute background noise to recent infections, leading to overestimation of incidence. Sensitivity analysis can be performed using a range of noise thresholds to assess robustness to potential misspecification of $\nu$.

Measurement noise, $\varepsilon$, represents laboratory assay variability from the laboratory testing process. Users of **serocalculator** should estimate $\varepsilon$ using the CV (coefficient of variation; i.e., the ratio of the standard deviation to the mean) for replicate measurements of the same biological samples.[2] Note that the CV should ideally be estimated using replicates across plates rather than within the same plate. Measurement noise can be positive or negative and can be reduced by running laboratory assays in duplicate and taking the mean.

Additional details for these noise parameters and their development were published by Teunis and Eijkeren in 2020 and specific noise estimation procedures were described by Aiemjoy *et al*. in 2022.[1,2]

### S7. Description of Enteric Fever Example Dataset

Enteric fever remains a major cause of morbidity and mortality in south Asia, southeast Asia, and sub-Saharan Africa, particularly for children.[1] It is caused by systemic infection with *Salmonella enterica* serovars Typhi and Paratyphi, which typically produce non-specific symptoms of fever, headache, and malaise. Complications can include intestinal perforation, gastrointestinal bleeding, altered mental state, septic shock, and death.[2,3] Although vaccines are available, many countries do not have the evidence of enteric fever burden to justify vaccine introduction. Estimates of seroincidence rates can provide the necessary data for vaccine introductions, while also specifying the population groups that may experience the highest risks of infection.

In the use case, we elect to estimate our seroincidence based on both IgA and IgG antibody responses to the Hemolysin E (HlyE) antigen (in column 'antigen_iso'), which have been shown to reliably distinguish enteric fever from other invasive bacterial infections.[4] Data for the use case comes from the SeroEpidemiology and Environmental Surveillance (SEES) for enteric fever study, which was conducted in Bangladesh, Nepal, and Pakistan.[5] It includes quantitative antibody responses from blood culture-confirmed enteric fever cases from the prospective clinical surveillance study, Surveillance for Enteric Fever in Asia Project (SEAP) that ran from 2016 to 2019 across sites in Bangladesh, Nepal, and Pakistan,[6] as well as population-based cross-sectional serologic samples collected between 2019-2021 from the same catchment areas.[5] The study sought to evaluate whether new diagnostic serological markers for enteric fever could reliably estimate population-level incidence.

In the population-based arm, households were selected using a single-stage, cluster random sampling method.[7] A grid was laid onto a map of each catchment area, dividing each area into 1000-2500 cells. Densely populated areas were further broken down into more cells of smaller size. Cells were then randomly selected from each catchment area and a field team member approached every household in that cell. Individuals were randomly selected using an age-stratified sample (age 0–4 years, age 5–9 years, age 10–15 years, and age 16–25 years). Participants were followed up to three times approximately 6 months apart.

The SEES study demonstrated that when combined with longitudinal antibody dynamics models, cross-sectionally collected antibody responses to HlyE and LPS could reliably estimate seroconversion rates and capture differences in seroconversion that correlated with clinical incidence estimates.[5] The available data from this study include quantitative antibody responses (IgA and IgG) to *S.* Typhi and *S.* Paratyphi antigens HlyE and LPS from both confirmed enteric fever patients and population-based participants from the same catchment areas. Full details of the SEAP and SEES studies have been published previously elsewhere.[5-7] The provided example does not factor in vaccination because the typhoid conjugate vaccine targets a different antigen, Vi (the virulence antigen), so it does not cross-react with the serological assays for HlyE and LPS.
